# High seroprevalence of *Leishmania infantum* is linked to immune activation in people with HIV: a two-stage cross-sectional study in Bahia, Brazil

**DOI:** 10.1101/2021.10.03.21264311

**Authors:** Laise de Moraes, Luciane Amorim Santos, Liã Bárbara Arruda, Maria da Purificação Pereira da Silva, Márcio de Oliveira Silva, José Adriano Góes Silva, André Ramos, Marcos Bastos dos Santos, Felipe Guimarães Torres, Cibele Orge, Antonio Marcos Teixeira, Thiago Vieira, Laura Ramírez, Manuel Soto, Maria Fernanda Rios Grassi, Isadora Cristina de Siqueira, Dorcas Lamounier Costa, Carlos Henrique Costa, Bruno de Bezerril Andrade, Kevan Akrami, Camila Indiani de Oliveira, Viviane Sampaio Boaventura, Manoel Barral-Netto, Aldina Barral, Anne-Mieke Vandamme, Johan Van Weyenbergh, Ricardo Khouri

## Abstract

**Background:** Visceral leishmaniasis is an opportunistic disease in HIV-1 infected individuals, although not yet recognized as a determining factor for AIDS diagnosis. The growing geographical overlap of HIV-1 and *Leishmania* infections is an emerging challenge worldwide, as co-infection increases morbidity and mortality for both. Here, we determined the prevalence of people living with HIV (PWH) with a previous or ongoing infection by *Leishmania infantum* in Bahia, Brazil and investigated the virological and immunological factors associated with co-infection.

**Methodology and Principal Findings:** We adopted a two-stage cross-sectional cohort (CSC) design (CSC-I, n=5,346 and CSC-II, n=317) of treatment-naïve HIV-1-infected individuals in Bahia, Brazil. In CSC-I, samples collected at the time of HIV-1 diagnosis between 1998 and 2013 were used for serological screening for leishmaniasis by an in-house immunoassay (ELISA) with SLA (Soluble Leishmania Antigen), resulting in a prevalence of previous or ongoing infection of 16.27%. Next, 317 PWH were prospectively recruited from July 2014 to December 2015 with collection of sociodemographic and clinical data. Serological validation by two different immunoassays confirmed a prevalence of 15.46% and 8.20% by anti-SLA, and anti-HSP70 serology, respectively, whereas 4.73% were double-positive (DP). Stratification of these 317 individuals in DP and double-negative (DN) revealed a significant reduction of CD4^+^ counts and CD4^+^/CD8^+^ ratios and a tendency of increased viral load in the DP group, as compared to DN. No statistical differences in HIV-1 subtype distribution were observed between the two groups. However, we found a significant increase of CXCL10/IP-10 (*p*=0.0076) and a tendency of increased CXCL9/MIG (*p*=0.061) in individuals with DP serology for *L. infantum*, demonstrating intensified immune activation in this group. These findings were corroborated at the transcriptome level in independent *Leishmania*- and HIV-1-infected cohorts (Swiss HIV Cohort and Piaui Northeast Brazil Cohort), indicating that *CXCL10* transcripts are shared by the IFN-dominated immune activation gene signatures of both pathogens and positively correlated to viral load in untreated PWH.

**Conclusions/Significance:** This study demonstrated a high prevalence of PWH with *L. infantum* seropositivity in Bahia, Brazil, linked to IFN-mediated immune activation and a significant decrease in CD4^+^ levels. Our results highlight the urgent need to increase awareness and define public health strategies for the management and prevention of HIV-1 and *L. infantum* co-infection.

**AUTHOR SUMMARY:** More than 1 billion people live in areas endemic for leishmaniasis and are at risk of infection, which includes one third of the 38 million people living with HIV (PWH) worldwide. Leishmaniasis is a neglected tropical disease caused by infection with *Leishmania* parasites, transmitted by bites from infected sand flies. HIV/*Leishmania* co-infection is increasing worldwide, especially in Southern Europe and Brazil, due to both human and ecological factors (migration, climate change, deforestation). This study demonstrates that a worryingly high number (one out of 6) of PWH in Bahia (Northeast Brazil) show previous or ongoing infection with the parasite *Leishmania infantum,* determined by antibodies detection against this parasite in PWH. Using a rigorous study design with data collection over 18 years in >5500 PWH, we found similarly high numbers (16.3% and 15.5%) of PWH with antibodies against *Leishmania infantum* in two consecutive cohorts. Moreover, PWH with antibodies against *Leishmania infantum* in Bahia, Brazil displayed a worse immune profile, with lower CD4 immune cells numbers and immune activation mediated by interferon, which we confirmed in independent HIV- and *Leishmania*-infected cohorts from other geographic areas. Our results underscore the need to raise awareness and define public health strategies to prevent HIV/*Leishmania* co-infection, since therapeutic failure and mortality are strongly increased in co-infected PWH, even with antiretroviral therapy as standard of care.

## INTRODUCTION

HIV is the etiological agent of Acquired Immunodeficiency Syndrome (AIDS), a slowly progressive disease characterized by chronic immune activation resulting from the loss of cell-mediated immune function [1, 2]. Host genetic factors (e. g. HLA genotypes) and host immune response (e.g. release of proinflammatory cytokines and chemokines), as well as the presence of co-infections, can directly influence the rate of disease progression [2–4].

Neglected Tropical Diseases (NTDs) are endemic among the most impoverished populations in Africa, Asia and Latin America. Among these NTDs, visceral leishmaniasis (VL), caused by infection with *Leishmania infantum*, is associated with increased morbidity and mortality in people with HIV-1 (PWH) but has not been recognized as a diagnostic criterion for AIDS [5]. The World Health Organization (WHO) identified the geographical overlap of both infections as an emerging challenge in successfully controlling the HIV-1 infection in countries considered endemic for leishmaniasis, such as Brazil [6].

The few studies evaluating the prevalence of HIV-1 and *L. infantum* coinfection on a national level in Brazil [7, 8] have highlighted regions endemic for leishmaniasis: Distrito Federal [9], Ceará [10, 11], Maranhão [12, 13], Mato Grosso [14], Mato Grosso do Sul [15, 16], Minas Gerais [17, 18], Pernambuco [19], Piauí [20], Rio Grande do Norte [21, 22], Sergipe [23] and Tocantins [24]. Bahia is also considered an endemic area for leishmaniasis, with an incidence rate of 0.08/10,000 inhabitants for VL reported in 2018 [25]. In addition, the increase of geographical overlap between HIV-1 and *Leishmania* infection in Bahia [25, 26] has raised public health concerns. However, no epidemiological studies have attempted to investigate the prevalence of co-infection to date. Therefore, we investigated the seroprevalence of *L. infantum* in PWH in the state of Bahia, Brazil and attempted to identify virological and immunological factors associated with co-infection, supported by publicly available transcriptomic data.

## METHODOLOGY

### Study population and design

A two-stage cross-sectional cohort study (CSC-I, 1998-2013 and CSC-II, 2014-2015) was designed to determine the seroprevalence of *L. infantum* in PWH and to evaluate the association with demographic, clinical, virological and immunological parameters (Figure 1). From 1998 to 2015, individuals with a new confirmed diagnosis of HIV-1 at the Specialized Center for Diagnosis, Care and Research (CEDAP), a state government public health reference service located in the city of Salvador, Bahia-Brazil, were included in this study. Their remaining blood sample was utilized for the outlined assays herein. First, serological reactivity to *L. infantum* was assessed in plasma or serum samples from the retrospective cross-sectional cohort (n=5,346) collected between 1998 and 2013. In the second stage, serological reactivity to *L. infantum* was assessed in a prospective cross-sectional cohort of 317 PWH (sampled between July 2014 and December 2015), with demographic and clinical data (sex, age, HIV-1 viral load, CD4^+^, CD8^+^, CD4/CD8 ratio and CD45^+^ T cell counts) prospectively obtained from the clinical records. Additionally, we performed a subpopulation analysis on a subset of PWH identified as either double-positive or double-negative using two independent anti-*Leishmania* immunoassays. Based on these stringently defined categories, we compared virological and immunological factors influenced by *L. infantum* seropositivity. The study’s recruitment did not interfere with standard medical care, and patients were treated according to the general Brazilian/WHO guidelines. This study was conducted following the Declaration of Helsinki and was approved by the Institutional Review Board of the Gonçalo Moniz Institute (IGM-FIOCRUZ) (protocol number 1.764.505).

**Figure 1.**
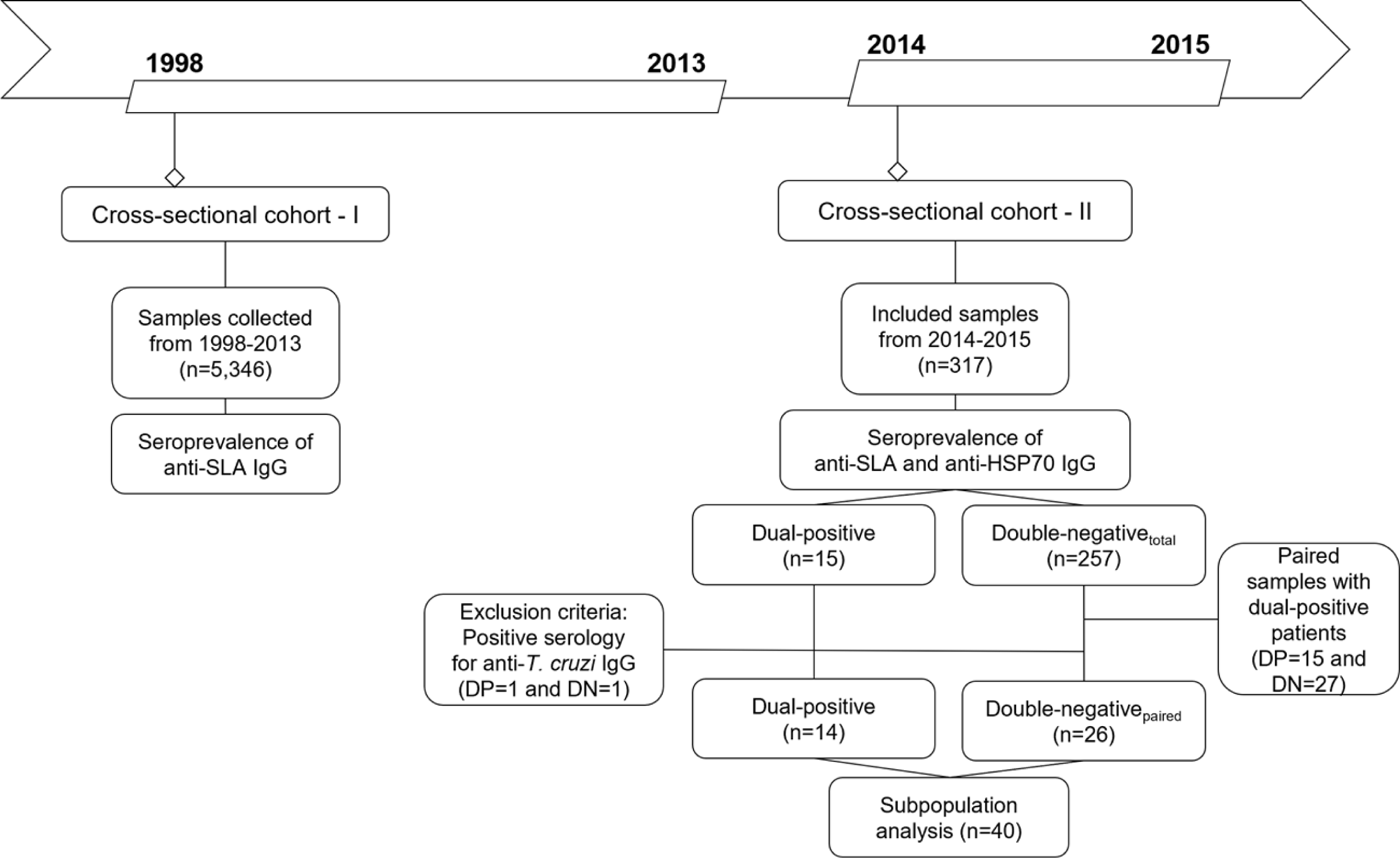
Flow chart of study design detailing the two-stage cross-sectional and prospective cohort study among people living with HIV in Bahia, Brazil.

### Detection of anti-*Leishmania and* anti-*Trypanosoma* antibodies

All samples underwent a previously described and validated in-house enzyme-linked immunosorbent assay (ELISA) for the detection of IgG against Soluble *Leishmania infantum* Antigen (SLA), which we previously demonstrated as a sensitive biomarker (90% sensitivity and 95% specificity) of current or past infection with the parasite [27]. SLA preparation and ELISA procedures were performed as previously described [28]. For the prospective cohort of 317 samples from PWH with digitalized clinical records, results of the SLA immunoassay were compared with a second, previously described ELISA employing recombinant *L. infantum* heat shock protein 70 (HSP70), for which we once found a sensitivity of 74% and a specificity of 73% [29]. To avoid false-positive results for *L. infantum* due to cross-reactivity among Trypanosomatidae parasites, an ELISA for anti-*Trypanosoma cruzi* IgG (Euroimmun) was employed in a subset of 42 samples with dual-positive (DP) (n=15) and double-negative (n=27) serology for anti-SLA and anti-HSP70.

### Control samples and ELISA cutoff value

Positive (n=6) *Leishmania-*infected patient samples and negative (n=47) control samples previously identified by the intradermal Leishmanin (also known as Montenegro) Delayed-type Hypersensitivity (DTH) skin test, with clinical and laboratorial confirmation, were used to establish anti-SLA and anti-HSP70 ELISA cutoff values. For each assay, specific cutoff values were determined using the mean OD values obtained from the included negative samples plus three times the observed standard deviation, with each assay’s index values corresponding to the OD/cutoff ratio (Supplementary Figure 1).

### Quantification of immunological markers

A Cytometric Bead Array (CBA) Human Inflammatory Cytokines Kit (BD Biosciences) was employed to quantify plasma concentrations of IL-1β, IL-6, IL-10, IL-12p70 and TNF, while CCL2/MPC-1, CCL5/RANTES, CXCL8/IL-8, CXCL9/MIG and CXCL10/IP-10 quantification was performed using a CBA Human Chemokine Kit (BD Biosciences), each following the manufacturer’s instructions. Data acquisition and analysis were performed using a FACSArray Bioanalyzer (BD Biosciences) and FlowJo v.10.0.5 (Tree Star) software, respectively.

### HIV-1 sequencing analyses

Viral RNA isolation was performed using a QIAamp Viral RNA Mini Kit (QIAGEN) following the manufacturer’s instructions. The protease/reverse transcriptase (PR/RT) region was amplified and sequenced as previously described [30]. Outer polymerase chain reaction (PCR) was performed using a SuperScript III One-Step RT-PCR System with Platinum Taq DNA Polymerase (Thermo Fisher Scientific) and the following primers: K1 (CAGAGCCAACAGCCCCACC) and K2 (TTTCCCCACTAACTTCTGTATGTCATTGACA) [31]. Inner PCR was performed using Platinum Taq DNA Polymerase High Fidelity (Thermo Fisher Scientific) with the following primers: DP16 (CCTCAAATCACTCTTTGGCAAC) and RT4 (AGTTCATAACCCATCCAAAG) [32]. The generated inner PCR products were sequenced using a BigDye Terminator v.3.1 Cycle Sequencing Kit (Applied Biosystems) with capillary electrophoresis run on an ABI 3500xL Genetic Analyzer (Applied Biosystems) employing the following primers:

F1 (GTTGACTCAGATTGGTTGCAC), F2 (GTATGTCATTGACAGTCCAGC [33], DP10 (CAACTCCCTCTCAGAAGCAGGAGCCG), DP11 (CCATTCCTGGCTTTAATTTTACTGGTA) [34], RT4 AGTTCATAACCCATCCAAAG), GABO1 (CTCARGACTTYTGGGAAGTTC) and GABO2 (GCATCHCCCACATCYAGTACTG) [30]. Sequence visualization, editing and assembly were performed using Geneious v.10.0.8 (Biomatters) software. HIV-1 subtyping was determined using the REGA HIV-1 Subtyping Tool v.3.4.1 (https://www.genomedetective.com/app/typingtool/hiv) and the jpHMM-HIV approach [35]. Phylogenetic analysis was conducted using HIV-1 subtype references retrieved from the Los Alamos HIV sequence database (https://www.hiv.lanl.gov/content/sequence/NEWALIGN/align.html) (Supplementary Table 1). Phylogenetic inference was performed using PhyML v.3.0 [36], applying the BIONJ method and using the General Time Reversible (GTR) nucleotide substitution model with 1,000 bootstrap replicates.

### Transcriptome and systems biology analysis

Transcriptome analysis of publicly available data was performed as recently described [37–39]. Overlap ratio was calculated as: k/K= # Genes in Overlap (k) / # Genes in Gene Set (K) using Molecular Signatures Database (MSigDb). HIV-1 (Swiss HIV Cohort, n=137) [40] and *Leishmania*-infected individuals (Piaui Northeast Brazil Cohort, n=30) [41] were analyzed using GEO2R from Gene Expression Omnibus (GEO).

### Statistical analysis

Since data were not normally distributed, as identified by the Shapiro-Wilk and D’Agostino-Pearson tests (GraphPad Prism v.7.0), results were analyzed using non-parametric tests: Mann-Whitney test and Spearman’s correlation, as well as Fisher’s exact test, with p<0.05 considered statistically significant.

## RESULTS

### Detection of IgG antibodies against *Leishmania (L.) infantum* antigens (anti-SLA and anti-HSP70) in people living with HIV (PWH)

To determine the seroprevalence to *L. infantum* among PWHs, 5,346 plasma or serum samples from treatment-näive HIV-1-infected individuals diagnosed between 1998 and 2013 were tested for the presence of IgG antibodies against soluble *L. infantum* antigen (anti-SLA) (Figure 1).

Positivity for anti-SLA serology was detected in 870/5,346 samples, resulting in an prevalence rate of 16.27% (Figure 2). Samples from patients with clinically confirmed diagnosis of visceral leishmaniasis (VL) and laboratory-confirmed *L. infantum* infection in the absence (n=16) or presence of HIV-1 co-infection (n=10) were used as an additional control [42]. As shown in Figure 2A, anti-SLA antibodies were detected in 81.25% and 100% of HIV-1-negative and HIV-1-positive VL patients, respectively. Besides, we found humoral immune response against SLA in VL patients quantitatively similar regardless of the presence of HIV-1 co-infection (Mann Whitney p=0.71, HIV-negative vs HIV-positive). Thus, we could confirm the high specificity of the *L. infantum* anti-SLA IgG serology protocol employed herein.

**Figure 2.**
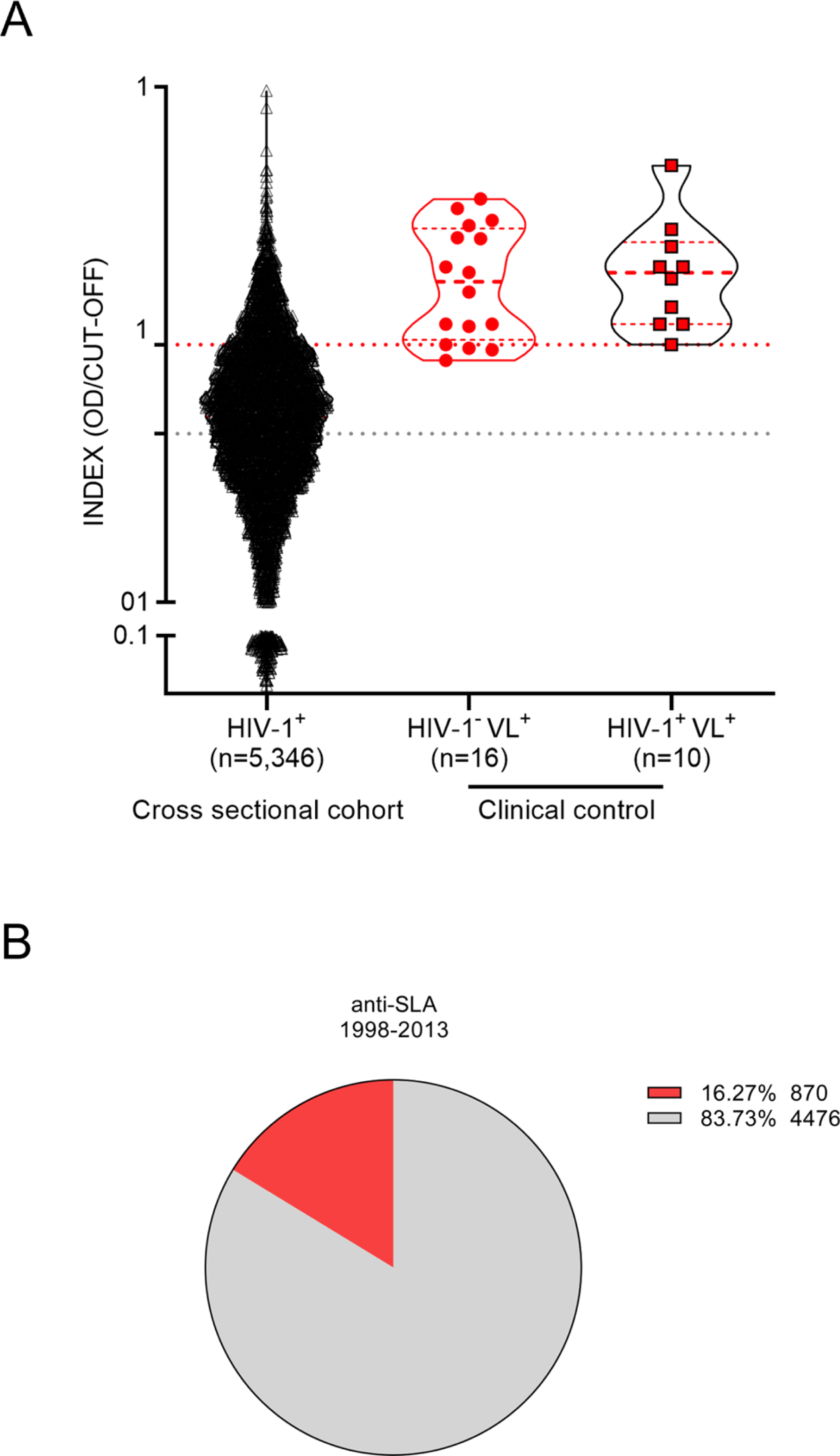
Detection of anti-*L. infantum* IgG antibodies in plasma/serum samples collected from PWH between 1998 and 2013. A) Violin plot with a dot-plot overlay of OD index values representative of *L. infantum* anti-SLA serology in the retrospective PWH cohort (n=5,346) and a clinical- and laboratory-confirmed control group (HIV-1^-^ VL^+^, n=16; and HIV-1^-^ VL^+^, n=10). Filled black circles (●): PWH cross-sectional cohort samples; filled red circles (●): clinical HIV-1 negative controls with VL; filled red squares with black borders (■): clinical controls with HIV-1 positive controls and VL. B) Pie chart representation of samples (expressed as percentages) with positive (red) and negative (gray) anti-SLA *L. infantum* serology.

Subsequently, to validate our findings, we enrolled an independent prospective CSC between 2014 and 2015, composed of 317 treatment-näive HIV-1-infected individuals from which complete demographic and clinical data were obtained from digitalized clinical records. Moreover, we combined anti-SLA serology with a second specific assay to detect IgG antibodies against *L. infantum* HSP70 recombinant protein (anti-HSP70) (Figure 1). The overall prevalence of anti-SLA positive serology in the prospective cohort was 15.46% (49/317), similar to the cross-sectional cohort results. The prevalence of anti-HSP70 positive serology in the prospective cohort was 8.20% (26/317) (Figure 3). Positivity for both anti-SLA and anti-HSP70 (i.e., double-positivity – DP) was detected in 4.73% (15/317) of the cohort; anti-SLA positive serology alone was detected in 10.73% (34/317); anti-HSP70 serology alone in only 3.47% (11/317) and the union of both anti-SLA and anti HSP70 positive serology was detected in 18.93% (Figure 3D).

**Figure 3.**
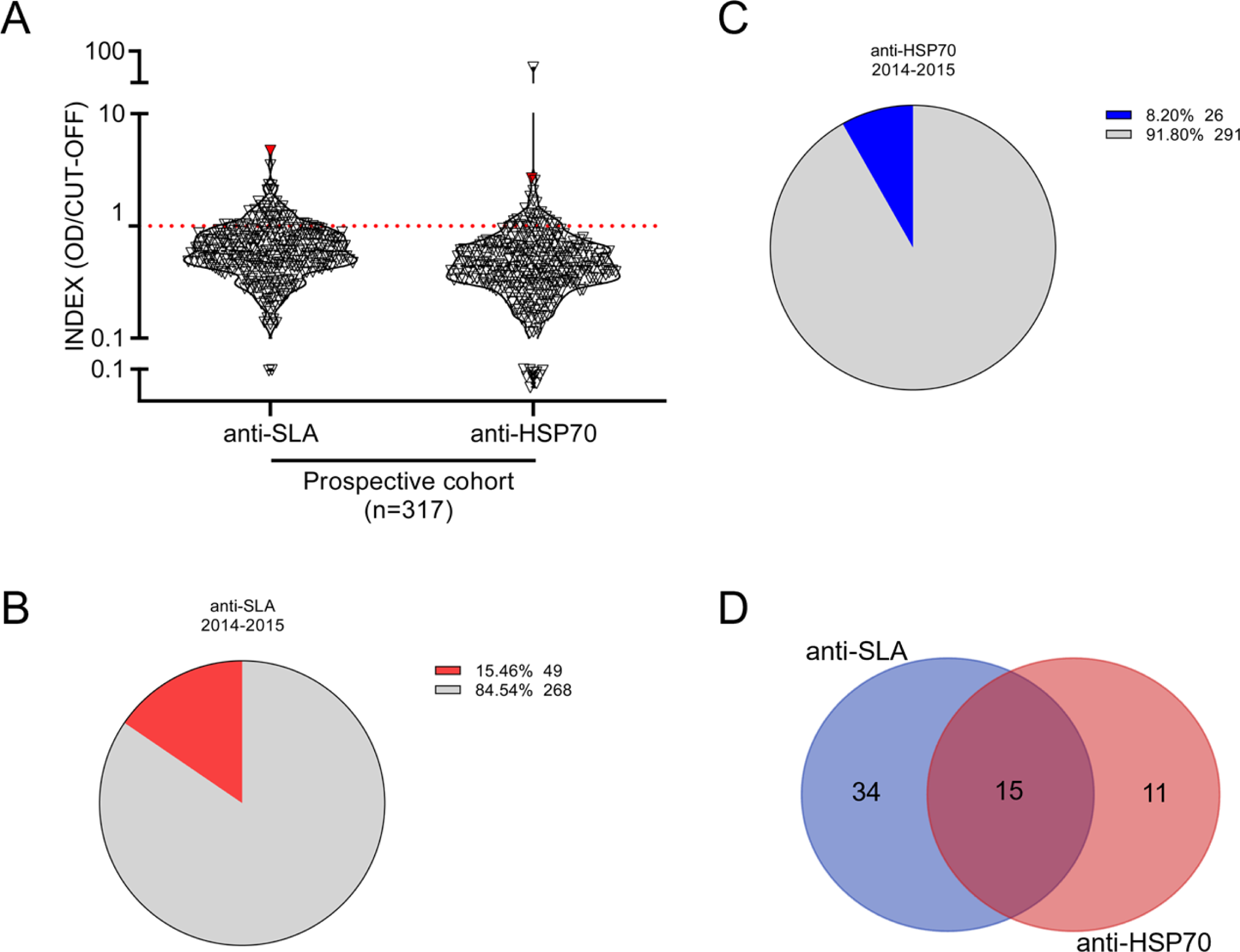
Detection of anti-*L. infantum* IgG antibodies in plasma samples collected from PWH between 2014 and 2015. A) Violin plot with a dot-plot overlay of OD index values representative of *L. infantum* anti-SLA and anti-HSP70 serology in the PWH prospective cohort (n=317). The red symbols represent the DP individual diagnosed with visceral leishmaniasis >20 years ago. B-C) Pie chart representation (expressed as percentages) of samples positive for B) *L. infantum* anti-HSP70 (blue) and C) *L. infantum* anti-SLA (red). Percentages of negative samples are depicted in gray. D) Venn diagram illustrating overlap in detection between *L. infantum* anti-SLA and anti-HSP70 serological methods.

Based on these findings, DP (n=15) was adopted as the most stringent criterium for *L. infantum* infection. Assessment of standard diagnosis of previous or ongoing visceral leishmaniasis (VL) documented in the clinical records showed that one of the 15 DP subjects had VL reported >20 years ago (anti-SLA index=4.71; anti-HSP70 index=2.67). After two decades of clinical diagnosis, this pronounced humoral response most likely reflects chronic antigen exposure, supporting the concept that *Leishmania* infection is lifelong in most patients as observed in murine models of disease [43–45]. These results, taken together, demonstrated that a significant number of PWH in the state of Bahia are unaware of their exposure to and/or infection by *L. infantum* and are at risk of developing new or relapsing visceral leishmaniasis.

### Demographic and clinical markers associated with dual-positive *L. infantum* serology in PWH

The CSC-II (n=317) consisted of predominantly young males (72.23%) with a median age of 32 years old at the time of HIV diagnosis, median viral load of 40,370 copies/mL (IQR=7,032-141,359), CD4^+^ T cell count of 369 cells/µL (IQR=163-609), CD8^+^ T cell count of 1,025 cells/µL (IQR=715-1,441), and CD45^+^ T cell count of 2,144 cells/µL (IQR=1,507-2,763) (Table 1).

**Table 1.** Epidemiological, virological and immunological characteristics of PWH in Bahia (Brazil) between 2014 and 2105, stratified according to DP and DN_total_ Data are expressed as median values, interquartile ranges (IIQ) and numbers of individuals evaluated (N), or as proportions (%) with the number of individuals evaluated in parentheses. The Mann-Whitney test was used to determine statistical differences between DP and DN_total_; *p* values considered statistically significant (p<0.05) are shown in bold. DP: dual-positive; DN_total_: total double-negative; NA: not analyzed

|  | n=317 | DP (n=15) | DN<sub>total</sub> (n=257) | *p* value |
| --- | --- | --- | --- | --- |
| Male, n | 72.23% (229) | 73.33% (11) | 70.42% (181) | NA |
| Age, years | 32 (26-41) | 41 (30-50) | 32 (25-39) | **0.0430** |
| HIV-1 viral load, copies/mL | 40,370 (7,032-141,359) | 64,746 (2,480-532,702) | 34,956 (6,330-127,747) | 0.1268 |
| CD4<sup>+</sup> T cell count, cell/μL | 369 (163-609) | 168 (107-449) | 405 (188-641) | **0.0208** |
| CD8<sup>+</sup> T cell count, cell/μL | 1,025 (715-1,441) | 950 (688-1,546) | 1025 (728-1,443) | 0.8679 |
| CD4<sup>+</sup>/CD8<sup>+</sup> T cell ratio | 0.33 (0.14-0.59) | 0.21 (0.060-0.37) | 0.35 (0.15-0.63) | **0.0282** |
| CD45<sup>+</sup> T cell count, cell/μL | 2,144 (1,507-2,763) | 1,758 (1,142-3,028) | 2,198 (1,585-2,793) | 0.3262 |

Comparative analyses between the DP (n=15) and total double-negative (DN_total_) (n=257) groups indicated that individuals with antibodies against both SLA and HSP70 *L. infantum* were older (p=0.0430), presented lower baseline CD4^+^ T cell counts (p=0.0208) and lower CD4^+^/CD8^+^ T cell ratios (p=0.0282), as well as a tendency towards higher HIV-1 viral load levels (p=0.1268) suggestive of an advanced stage of HIV-1 infection than individuals without *L. infantum* antibodies (Table 1). Indeed, a large proportion of DP individuals (10/15, 66.67%) had <200 CD4+ T cells/µL at the time of HIV-1 diagnosis, which is considered AIDS-defining.

### Subpopulation analysis reveals virological and immunological markers associated with dual-positive *L. infantum* serology in PWH

To ensure specificity for *L. infantum* in the DP serological samples and to exclude the possibility of cross-reactivity due to polyclonal activation and hypergammaglobulinemia, anti-*Trypanosoma cruzi* IgG serology was performed. Samples demonstrating positivity for anti-*T. cruzi* IgG (n=2) were excluded from subsequent analyses (Supplementary Figure 2). Next, we performed a subpopulation analysis by matching DN individuals to the DP group by gender, age, viral load and CD4^+^ T cell count. This strategy resulted in the establishment of a third group (DN_paired_) (Figure 1), thereby mitigating the possibility of bias arising from polyclonal activation, hypergammaglobulinemia, high viral load or low CD4^+^ T cell levels. Controlling for these parameters limited the potential for variations in the HIV-1 transcriptomic profile and the state of immune activation. No statistical differences were observed between the DP and DN_paired_ groups regarding sociodemographic and clinical parameters, which underscores the pairing strategy’s equity (Table 2).

**Table 2.** Epidemiological, virological and immunological characteristics of PWH in Bahia (Brazil) between 2014 and 2105, stratified according to DP and DN_paired_. Data are expressed as median values, interquartile ranges (IIQ) and numbers of individuals evaluated (N), or as proportions (%) with the number of individuals evaluated in parentheses. The Mann-Whitney test was used to determine statistical differences between DP and DN_paired_; *p* values considered statistically significant (p<0.05) are shown in bold. DP: dual-positive; DN_paired_: total double-negative; NA: not analyzed

|  | DP (n=14) | DN<sub>paired</sub> (n=26) | *p* value |
| --- | --- | --- | --- |
| Male, n | 78.57% (11) | 88.46% (23) | NA |
| Age, years | 42 (29-51) | 33 (26-43) | 0.2224 |
| HIV-1 viral load, copies/ml | 147,580 (2,284-700,074) | 76,298 (15,757-141,159) | 0.4572 |
| CD4+ T cell count, cell/μl | 166 (91-450) | 300 (128-472) | 0.4113 |
| CD8+ T cell count, cell/μl | 1,008 (744-1,607) | 995 (852-1,489) | 0.7474 |
| CD4+/CD8+ T cell ratio | 0.21 (0.058-0.39) | 0.29 (0.14-0.49) | 0.3954 |
| CD45+ T cell count, cell/μl | 1,788 (1,262-3,054) | 1,958 (1,628-2,585) | 0.6641 |

To investigate possible associations between transmission clusters of a more-aggressive HIV subtype and worsen HIV clinical status, HIV-1 subtyping was performed in the DP and DN_paired_ subset to infer. After excluding samples with a viral load <1,000 copies/mL, both groups were pooled together (n=19) and submitted to phylogenetic analysis using the jpHMM-HIV model (Supplementary Figure 3). Concerning subtype distribution, subtype B (63.16%, 12/19) was the most prevalent, followed by subtypes C (15.79%, 3/19), recombinant BF (15.79%, 3/19) and D (5.26%, 1/19). No statistical differences in subtype were observed between the DP and DN_paired_ groups (p=0.67) (Supplementary Table 2). No transmission cluster was identified. HIV-1 subtyping showed no impact on *L. infantum* serology titers, viral load, T cell count, or any other host immunological marker evaluated, indicating that HIV-1 subtypes B, C, D, F1 and BF exhibit similar virological and immunological outcome in HIV/*L. infantum* co-infection.

Next, we quantified circulating cytokines and chemokines that might reflect an increase in systemic immune activation and have been previously associated with infection to *L. infantum* [46–49]. The systemic levels of cytokines tested (IL-1β, IL-6, IL-10, IL-12p70 and TNF) were below the detection limits in all samples (DP and DN_paired_ groups), concordant with our previous findings in a large Cuban cohort of PWH [4]. Conversely, most chemokines (except IL-8) were readily detected in most PWH, demonstrating a significant increase in CXCL10/IP-10 (p=0.0076) and a tendency towards higher CXCL9/MIG levels (p=0.061) (Figure 4) in the DP group compared to DN_paired_, suggesting increased immune activation in the group with DP serology for *L. infantum*.

**Figure 4.**
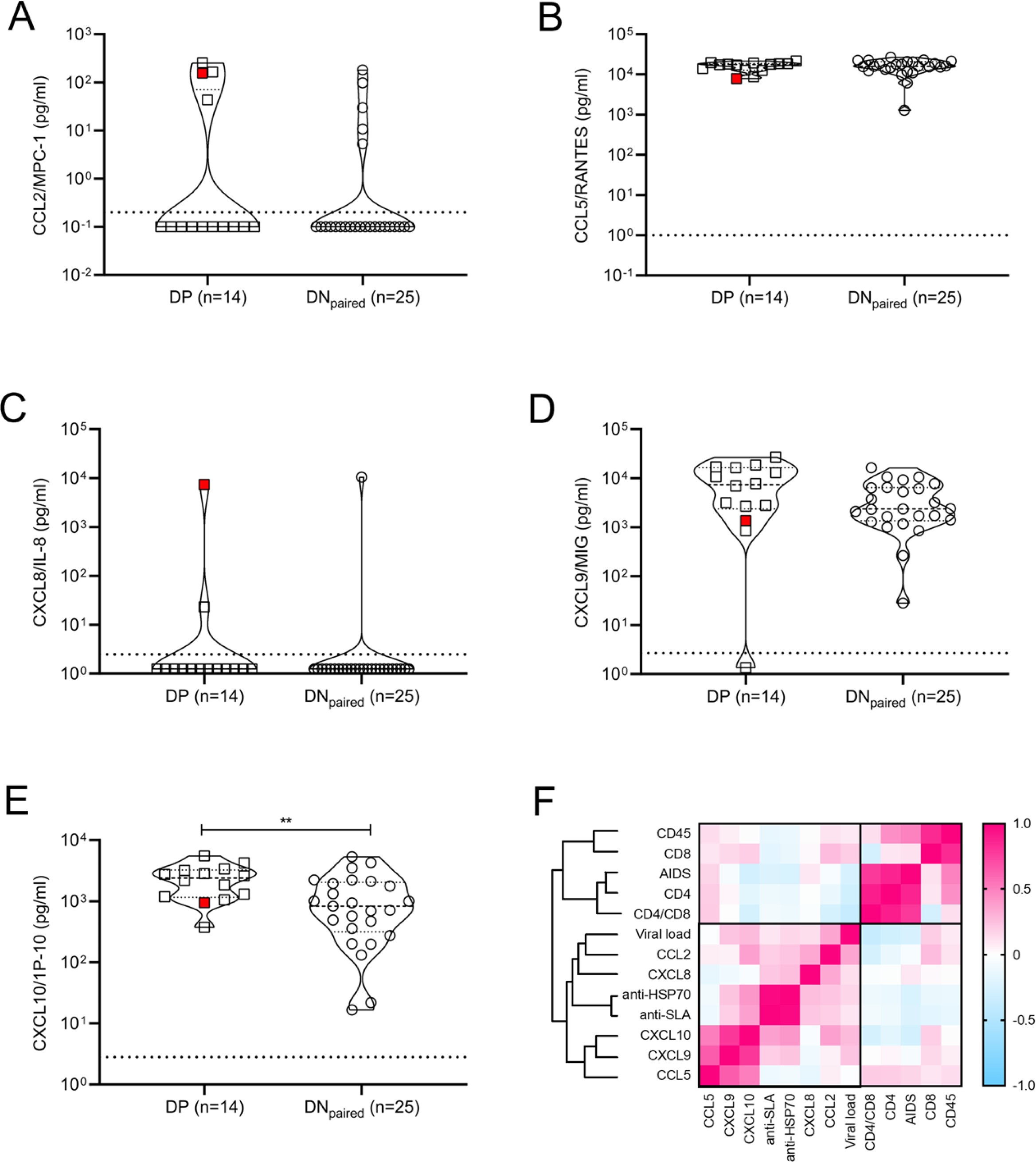
Chemokine profiles obtained from plasma/serum samples of DP and DN_paired_ groups. A-E) Violin plots with dot-plot overlays illustrating chemokine plasma levels (pg/mL). Statistical comparisons made by the Mann-Whitney test in plasma levels (pg/mL) of CCL2/MPC-1 (p=0.43), CCL5/RANTES (p=0.94), CXCL8/IL-8 (p=0.54), CXCL9/MIG (p=0.061) and CXCL10/IP-10 (p=0.0076) chemokines. Open squares (☐): subgroup of double-positive PWH (positive for both anti-SLA and anti-HSP70 *L. infantum* antigens); open circle (○): subgroup of paired double-negative PWH (serologically negative for anti-SLA and anti-HSP70 *L. infantum* antigens). The red symbol represents the DP individual diagnosed with visceral leishmaniasis >20 years ago. F) Heat map shows the correlation (pink: positive correlation; blue: negative correlation; white: no correlation) of epidemiological, virological and immunological characteristics of PWH.

This finding, together with the decreased CD4^+^ T cell counts found in our previous analysis (Table 2), corroborates the hypothesis that therapy-naïve HIV-1-infected subjects previously or currently infected with *L. infantum* exhibit a more intensified state of immune activation than HIV-1 positive, *L. infantum* seronegative subjects. During clinical follow-up, the introduction of antiretroviral therapy restored CD4^+^ T cell counts (median values of CD4^+^ T cell counts after 4-5 years of follow-up: DP=620 cells/µL and DN_paired_=870 cells/µL) and reduced viral load levels to undetectable limits similarly in each group (Figure 5), with no statistically significant differences between groups (AUC comparison, data not shown).

**Figure 5.**
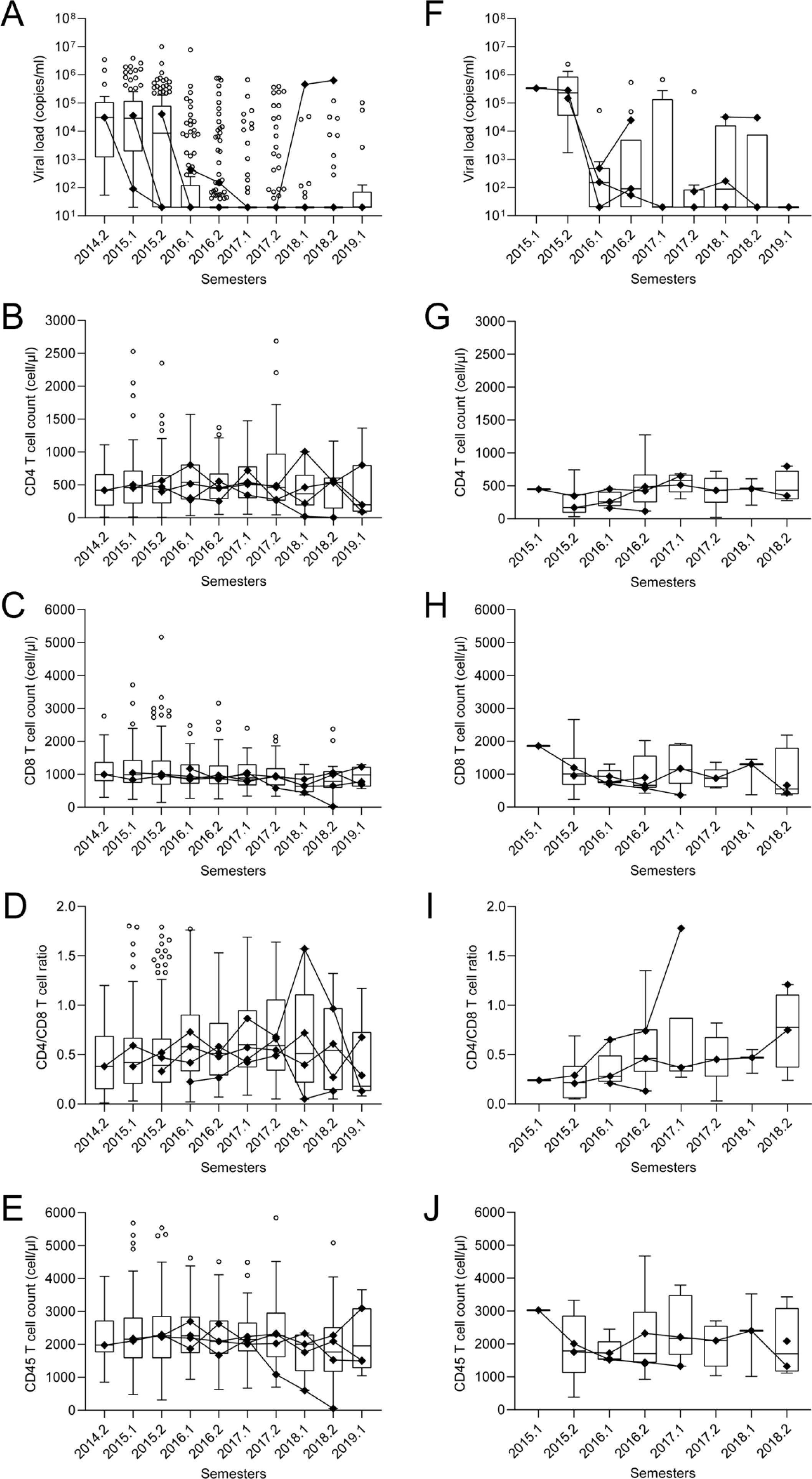
Long-term follow-up of CD4^+^, CD8^+^, CD4/CD8 ratio, CD45^+^ and viral load levels after therapy introduction in DP and DN_paired_ groups. A-J) Box plots with line plot overlays connecting medians of CD4^+^, CD8^+^, CD4/CD8 ratio, CD45^+^ and viral load levels during follow-up. A and F) Follow-up of viral load (copies/mL) in DN_paired_ (A) and DP (F) groups. B and G) Follow-up of CD4^+^ T cell count (cells/µL) in DN_paired_ (B) and DP (G) groups. C and H) Follow-up of CD8^+^ T cell count (cells/µL) in DN_paired_ (C) and DP (H) groups. D and I) Follow-up of CD4/CD8 (ratio) in DN_paired_ (D) and DP (I) groups. E and J) Follow-up of CD45^+^ T cell count (cells/µL) in DN_paired_ (E) and DP (J) groups.

### *CXCL10* is shared between the HIV and visceral leishmaniasis gene signatures, while *CXCL10, CXCL9* and *CCL2* positively correlate with HIV viral load in untreated PWH

Given the small number of immune activation markers and modest cohort size for the paired DP and DN groups, we aimed to validate and extend our findings by cross-examining independent cohorts of HIV-1 (Swiss HIV Cohort, n=137) or *Leishmania*-infected individuals with and without active disease (Piaui Northeast Brazil, n=30) with publicly available transcriptome-wide data [40, 41]. As shown in Figure 6A-B, a significant overlap (41 genes, enrichment p<0.0001) was found between the HIV and visceral leishmaniasis gene signatures when comparing the top 500 transcripts significantly upregulated during active disease in both untreated HIV patients and untreated visceral leishmaniasis patients, as compared to their paired post-treatment groups. A systems biology analysis of the 41 genes shared by HIV and leishmaniasis active disease signatures revealed several significantly enriched biological processes, encompassing immune activation broadly and, more specifically, type I IFN signaling and antiviral response. The overlapping genes included *CXCL10*, which we found to be overexpressed in the DP group of our prospective cohort, as well as *STAT1*, the major transcription factor activated by IFN signaling. As detailed in Figure 6A and Supplementary Tables 3-5, enriched gene sets from Molecular Signatures Database (MSigDb) representing ’immune activation’ included graft-versus-host disease, antigen recognition and blister cytotoxicity, while enriched gene sets representing ’type I IFN/antiviral response’ comprised those upregulated in the liver by HBV infection, by IFN-alpha treatment of fibroblasts *in vitro* and by IFN-beta treatment of multiple sclerosis patients *in vivo*. Of interest, *IFNG* transcripts significantly decrease in monoinfected visceral leishmaniasis individuals, confirming our previous results obtained at the protein level for systemic IFN-γ in two independent VL cohorts [46, 49].

**Figure 6.**
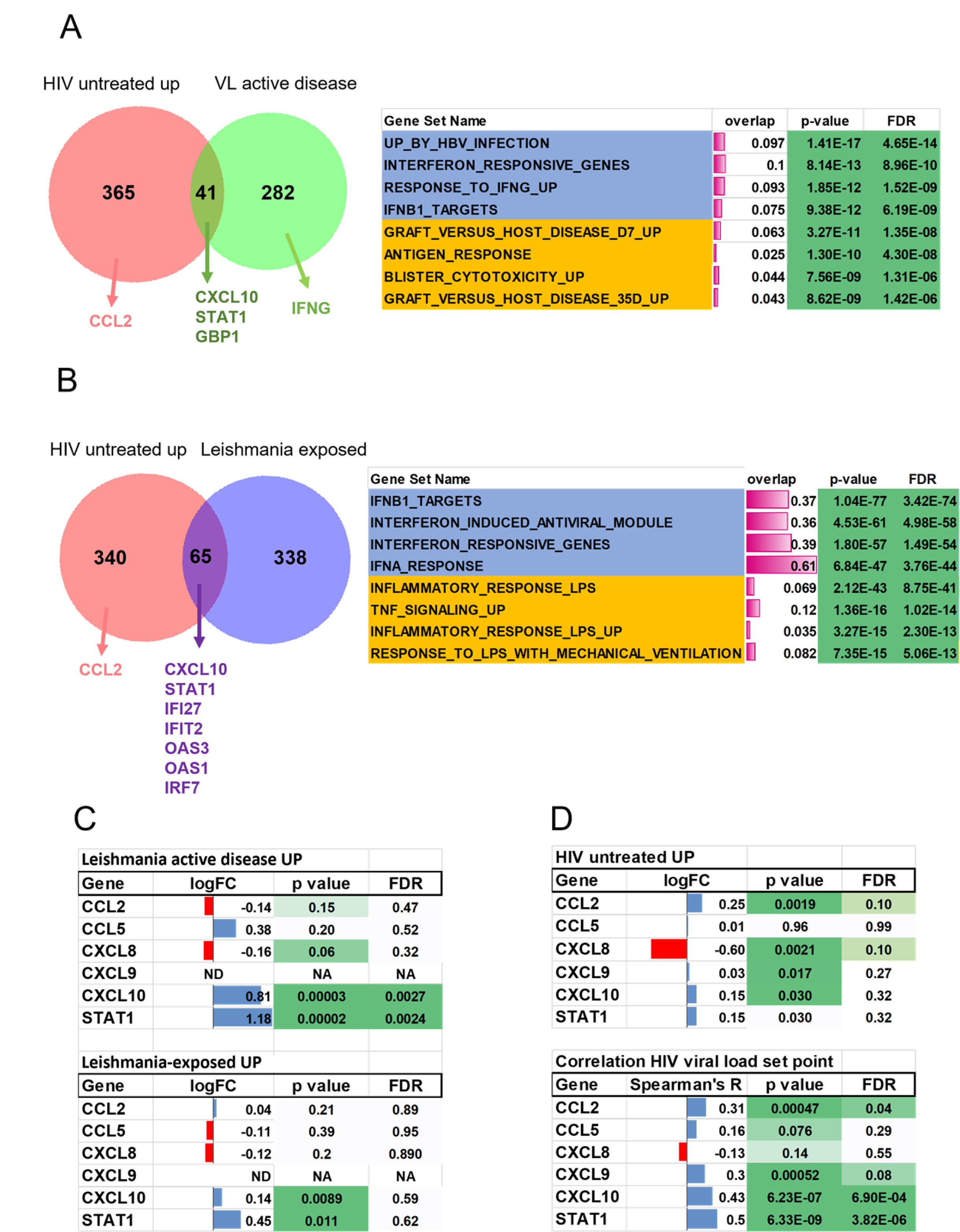
CXCL10 is shared between the HIV and visceral leishmaniasis disease signatures while *CXCL10, CXCL9* and *CCL2* transcripts positively correlate with HIV viral load in untreated PWH. Venn diagrams show overlap between (A) active disease gene signatures, i.e. genes upregulated in untreated PWH (Swiss cohorts) and visceral leishmaniasis patients (as compared to post-treatment) and (B) genes upregulated in untreated PWH and asymptomatic *Leishmania*-infected individuals (who did not develop disease). Systems biology analysis shows a significant enrichment of gene sets comprising immune activation and type I IFN signaling (right-hand panels). (C) Selective up-regulation of *CXCL10* and *STAT1* (as a positive control for IFN signaling) transcripts in both untreated visceral leishmaniasis patients (upper panel) and *Leishmania infantum*-exposed individuals (D) Selective up-regulation (upper panel) and positive correlation to viral load (lower panel) of *CXCL10, CXCL9 and CCL2* and *STAT1* (as a positive control for IFN signaling) transcripts in untreated PWH (Swiss cohort, n=137).

To define a specific ’*Leishmania* infection’ gene signature, we used the top 500 significantly upregulated transcripts from *Leishmania*-infected individuals who did not develop disease, for which *Leishmania* infection was documented by a positive skin DTH reaction, as compared to matched DTH-negative individuals from the same endemic area [41]. Surprisingly, the overlap between the ’untreated HIV’ and ’*Leishmania*-seropositive (DP)’ gene signatures was even more pronounced than the overlap between the ’untreated HIV’ and ’untreated visceral leishmaniasis’ gene signatures, both by quantitative (65 shared genes, Figure 6B) and qualitative measures (median gene set overlap 9.4% vs 3.2%, Mann-Whitney p<0.0001, Supplementary Table 3), indicating that even asymptomatic *Leishmania* infection has a profound effect on the systemic immune response. Again, *CXCL10* and *STAT1* were shared among the two disease signatures (Figure 6B), and enriched gene sets included type I IFN signaling (genes upregulated by IFN-beta treatment of multiple sclerosis patients *in vivo* and by IFN-alpha treatment of fibroblasts *in vitro*) and immune activation (inflammatory response to LPS, TNF signaling).

Validation of our DP cohort results revealed that *CXCL10*, but not *CCL2, CCL5* or *CXCL8* transcripts, were significantly upregulated by *Leishmania infantum* infection with or without clinically definite visceral leishmaniasis (Figure 6C upper and lower panel). In the Swiss HIV Cohort, we found that *CXCL10*, *CCL2* and, to a lesser extent, *CXCL9* transcripts were significantly upregulated during untreated HIV-1 infection. In contrast, no significant effect was observed for *CXCL8* in either asymptomatic or symptomatic *Leishmania infantum* infection, supporting our negative findings for *CXCL8* genetic variation in visceral leishmaniasis patients from the same endemic region [47]. However, *CXCL8* transcripts were significantly downregulated during untreated HIV-1 infection (Figure 6D upper panel). Finally, as shown in Figure 6D (lower panel), *CCL2*, *CXCL9* and *CXCL10* transcripts were significantly correlated (even after stringent correction for genome-wide testing) with set point viral load in untreated PWH from the Swiss HIV Cohort (n=137), confirming the results from our prospective cross-sectional cohort (Figure 4F).

## DISCUSSION

The geographic overlap of HIV-1 and *L. infantum* infections raises the possibility of co-infection, which several groups have shown to increase morbidity and mortality [7,11,52–58,13,14,19,21–23,50,51]. Several groups have studied the extent of this overlap in several Brazilian regions. However, no studies have attempted to determine the seroprevalence of *L. infantum* among PWH in the state of Bahia, despite the significant prevalence of both infections [25, 26]. We used a two-stage cross-sectional cohort approach (CSC-I, n=5,346 and CSC-II, n=317) and identified positive serology for *L. infantum* ranging from 4.73% (DP for anti-SLA and anti-HSP70) to 18.93% in the CSC-II (union of all positive samples for anti-SLA and anti-HSP70), with an intermediate seroprevalence for only anti-SLA of 16.27% in the CSC-I and 15.46% in the CSC-II. The observed frequencies of *Leishmania* seroprevalence in PWH using any of the serology strategies in both the cross-sectional cohorts support previous findings falling within the range of what has already been described in other Brazilian regions [9,12,20,59]. Moreover, demographic data indicated that all DP patients were born in the macroregions surrounding the cities of Salvador and Feira de Santana, which are responsible for 33.8% (1,074/3,172) of notified VL cases in Bahia between 2007 and 2015 [25].

The observed discrepancy between the results obtained with the two different immunoassays might be explained by the rigor employed to determine cutoffs for each serological test and the imbalance in protein fractions between these two antigen-based diagnostic methods. SLA is a heterogeneous mix of antigens and has a lower concentration of HSP70 than parasite structural proteins (actin, tubulin, etc.). Even though it will pick up a broader set of samples, some samples with low antibody levels will fail. On the other hand, the recombinant HSP70 antigen is homogenous and highly concentrated and has a better detection limit. We considered only those samples demonstrating dual positivity to increase confidence regarding past or present *L. infantum* infection. This strategy was underscored by the identification of one individual with a previous history of VL in the DP group and no cases with an earlier history of VL in the DN_paired_ group.

Unfortunately, only serum or plasma samples were available for HIV-1 RNA extraction, thus precluding DNA analysis to detect *L. infantum* by qPCR, which we acknowledge as a limitation of our study. However, Fukutani *et al.* (2014) [60] demonstrated a concordance of 68% between anti-SLA serology and PCR, and found a 5.40% rate of ongoing asymptomatic *L. infantum* infection among blood donors in Salvador, Bahia. Nevertheless, positive IgG serology for *L. infantum* indicates immunological memory of at least a previous *Leishmania* infection before HIV diagnosis. Strikingly, we identified a DP individual with a pronounced humoral response two decades after a VL clinical diagnosis. This ongoing humoral response most likely reflects chronic antigen exposure, emphasizing the concept that *Leishmania* infection is lifelong in most patients, as well as murine models of disease [43–45]. A wealth of studies worldwide [14,42,61–64] and a recent systematic review [56] have demonstrated a high frequency of relapse of VL in PWH, as well as increased mortality. Relapse of VL have been reported after anti-leishmaniasis treatment, even years after the patient was considered clinically cured, especially in PWH and/or immunosuppressive conditions [13, 51].

Prior large studies, including from our group, have found striking differences in the prevalence of VL within different regions in Brazil, as well as Southern Europe. The highest seroprevalence was observed in Mato Grosso, 41.40% of blood donors, in contrast to other regions in Brazil (Salvador: 5.40% of 700 [60]; Paraná: 11.40% of 176 [65]; Fortaleza: 17.10% of 431 [66]), and in Southern Europe (France: 13.40% of 565 [67]; Spain: 3.10% of 1,437 [68]).

Studies conducted in populations of newly diagnosed HIV-1-infected individuals from regions endemic for leishmaniasis in Brazil reported a prevalence of HIV-1/*L. infantum* co-infection ranging from 4.20% to 20.20%. In these studies, *L. infantum* infection was determined by parasitological, serological and/or molecular testing [9,12,20,59]. However, in co-infection studies focused on VL patients, which were diagnosed by parasitological, molecular or immunofluorescence methods, the prevalence of HIV-1/*L. infantum* coinfection ranged from 36.60% to 55.60% [16–18].

The present study also investigated possible differences in the prevalence of HIV-1 variants associated with *L. infantum* infection in PWH. With these subtyping results, we would like to investigate if a possible worsen disease progression in PWH with positive serology for *L. infantum* cases was related to cluster of transmission of a more-aggressive HIV subtype (e.g. HIV Subtype D) [69]. However, no differences were found concerning HIV-1 isolates between matched samples of patients infected (DP) or not (DN_paired_) to *L. infantum*. Within these isolates, subtype B (63.16%) was found to be the most prevalent, followed by C (15.79%), recombinant BF (15.79%) and D (5.26%). Recent studies in Bahia that evaluated HIV-1 variant distribution have described the wide distribution of subtype B, followed by a smaller proportion of recombinant BF units [70–72]. Of note, the higher prevalence of subtype C (15.79%) found herein stands in contrast to other studies also conducted in this region, which reported a very low prevalence ranging from 1.70% to 2.50% [71,73,74].

Moreover, our confirmation of the circulation of subtype D in the state is following work by Monteiro *et al*. (2009) [75], who previously reported the presence of subtype D in individuals from Feira de Santana (0.60% of _pol_F/_env_D subtype), a city located ∼100 km from Salvador. It is worth noting that the subtyping analysis performed herein involved three different methodologies (REGA HIV-1 Subtyping Tool, phylogeny and jpHMM-HIV probabilistic modeling), which lends support to the presently reported distribution. Furthermore, our subtype identification was performed using HIV-1 pol sequences, inferring subtype frequencies can vary when complete genome sequencing is performed.

Prior research into host immune responses has demonstrated that *L. infantum* infection induces a higher level of immune activation in co-infected PWH, which could lead to a more rapid progression to AIDS [3,76–78]. Our study found a roughly five-fold increase in circulating CXCL10/IP-10 and CXCL9/MIG chemokines in PWH exposed to *L. infantum,* supporting a link between positive serology for *L. infantum* and immune activation, which we validated at the transcript level for CXCL10 in independent HIV-1 and *Leishmania*-infected cohorts. These chemokines are strongly induced by both type I (IFN-α/β) and type II IFN (IFN-γ) [79–82]. It has been previously reported that Th1 subpopulations of CD4^+^CD95^+^ T cells in HIV-1 infected individuals are highly susceptible to HIV-1 envelope gp120-induced apoptosis [83, 84], which may corroborate the deleterious role of intensified immune activation [85–87]. In a submitted manuscript (Khouri *et al*.), we demonstrated an increase in both systemic soluble Fas as well as membrane-bound Fas/CD95 expression in CD8^+^ cells in HIV/VL co-infection, as compared to HIV-1 or VL mono-infected groups, in a large cohort of individuals recruited during an outbreak of VL in Piaui, another state in the same northeast region of Brazil.

Since we previously demonstrated STAT1-mediated IFN signaling as a preferential inducer of Fas/CD95 at both the protein and transcript level [88, 89], this study as well as Khouri *et al*. (submitted) underscore the pivotal role of IFN-driven immune activation in HIV-1 and *Leishmania infantum* co-infection and its deleterious effect on both AIDS progression and visceral leishmaniasis.

The present study aimed to describe the prevalence and consequences of *L. infantum* infection in a population of antiretroviral treatment-naïve PWH at diagnosis. We found a high seroprevalence of *L. infantum* in treatment-naïve PWH in Bahia (Brazil) associated with a more intensified state of immune activation as compared to HIV-1 monoinfected subjects, possibly exacerbating progression to AIDS, which was observed in 66.67% in co-infected individuals at diagnosis. Moreover, *Leishmania*-infected PWH are at high risk of developing severe visceral leishmaniasis [56]. These findings underscore the urgent need to increase awareness and define public health strategies for the management and prevention of HIV-1 and *L. infantum* co-infection.

## Data Availability

The data supporting the findings of this study are available within the article and its supplementary materials.

## FUNDING

This work was financed in part by Fundação de Amparo à Pesquisa do Estado da Bahia (FAPESB, APP0032/2016), Coordenação de Aperfeiçoamento de Pessoal de Nível Superior (CAPES – finance code 001) and FWO (grant G0D6817N). None of the funding organizations had any role in the study design, data collection, data interpretation or writing of this report.

## ACKNOWLEDGMENTS

The authors would like to thank Andris K. Walter for English language revision and manuscript copyediting assistance.

## Notes

### Competing Interest Statement

The authors have declared no competing interest.

### Author Declarations

This study was conducted following the Declaration of Helsinki and was approved by the Institutional Review Board of the Gonçalo Moniz Institute (IGM-FIOCRUZ) (protocol number 1.764.505).

## REFERENCES

1. Costin JM. Cytopathic Mechanisms of HIV-1. Virol J. 2007;4: 100. doi:10.1186/1743-422X-4-100

2. Perkins MR, Bartha I, Timmer JK, Liebner JC, Wollinsky D, Günthard HF, et al. The Interplay Between Host Genetic Variation, Viral Replication, and Microbial Translocation in Untreated HIV-Infected Individuals. J Infect Dis. 2015;212: 578–584. doi:10.1093/infdis/jiv089

3. Claiborne DT, Prince JL, Scully E, Macharia G, Micci L, Lawson B, et al. Replicative fitness of transmitted HIV-1 drives acute immune activation, proviral load in memory CD4 + T cells, and disease progression. Proc Natl Acad Sci. 2015;112: E1480–E1489. doi:10.1073/pnas.1421607112

4. Kouri V, Khouri R, Alemán Y, Abrahantes Y, Vercauteren J, Pineda-Peña A-C, et al. CRF19_cpx is an Evolutionary fit HIV-1 Variant Strongly Associated With Rapid Progression to AIDS in Cuba. EBioMedicine. 2015;2: 244–254. doi:10.1016/j.ebiom.2015.01.015

5. van Griensven J, Ritmeijer K, Lynen L, Diro E. Visceral Leishmaniasis as an AIDS Defining Condition: Towards Consistency across WHO Guidelines. Ghedin E, editor. PLoS Negl Trop Dis. 2014;8: e2916. doi:10.1371/journal.pntd.0002916

6. World Health Organization. Investing To Overcome the Global Impact of Neglected Tropical Diseases: third WHO Report on Neglected Tropical Diseases. 2015.

7. Coura-Vital W, Araújo VEM de, Reis IA, Amancio FF, Reis AB, Carneiro M. Prognostic Factors and Scoring System for Death from Visceral Leishmaniasis: An Historical Cohort Study in Brazil. Santiago H da C, editor. PLoS Negl Trop Dis. 2014;8: e3374. doi:10.1371/journal.pntd.0003374

8. Leite de Sousa-Gomes M, Nilce Silveira Maia-Elkhoury A, Maria Pelissari D, Edilson Ferreira de Lima Junior F, Martins de Sena J, Paula Cechinel M. Coinfecção Leishmania-HIV no Brasil: aspectos epidemiológicos, clínicos e laboratoriais. Epidemiol e Serviços Saúde. 2011;20: 519–526. doi:10.5123/S1679-49742011000400011

9. Carranza-Tamayo CO, Assis TSM de, Neri ATB, Cupolillo E, Rabello A, Romero GAS. Prevalence of Leishmania infection in adult HIV/AIDS patients treated in a tertiary-level care center in Brasilia, Federal District, Brazil. Trans R Soc Trop Med Hyg. 2009;103: 743–748. doi:10.1016/j.trstmh.2009.01.014

10. Távora LGF, Nogueira MB, Gomes ST. Visceral Leishmaniasis/HIV co-infection in northeast Brazil: evaluation of outcome. Brazilian J Infect Dis. 2015;19: 651– 656. doi:10.1016/j.bjid.2015.07.004

11. Diniz LFB, de Souza CDF, do Carmo RF. Epidemiology of human visceral leishmaniasis in the urban centers of the lower-middle São Francisco Valley, Brazilian semiarid region. Rev Soc Bras Med Trop. 2018;51: 461–466. doi:10.1590/0037-8682-0074-2018

12. Carvalho FL, Aires DLS, Segunda ZF, Azevedo CMPES De, Corrêa RDGCF, Aquino DMC de, et al. Perfil epidemiológico dos indivíduos HIV positivo e coinfecção HIV-Leishmania em um serviço de referência em São Luís, MA, Brasil. Cien Saude Colet. 2013;18: 1305–1312. doi:10.1590/S1413-81232013000500015

13. Silva de Lima UR, Vanolli L, Moraes EC, Ithamar JS, Pedrozo e Silva de Azevedo C de M. Visceral leishmaniasis in Northeast Brazil: What is the impact of HIV on this protozoan infection? PLoS One. 2019;14: 1–14. doi:10.1371/journal.pone.0225875

14. Luz JGG, Naves DB, de Carvalho AG, Meira GA, Dias JVL, Fontes CJF. Visceral leishmaniasis in a Brazilian endemic area: An overview of occurrence, HIV coinfection and lethality. Rev Inst Med Trop Sao Paulo. 2018;60: 1–9. doi:10.1590/S1678-9946201860012

15. Botelho ACA, Natal D. Primeira descrição epidemiológica da leishmaniose visceral em Campo Grande, Estado de Mato Grosso do Sul. Rev Soc Bras Med Trop. 2009;42: 503–508. doi:10.1590/S0037-86822009000500006

16. Druzian AF, Souza AS de, Campos DN de, Croda J, Higa MG, Dorval MEC, et al. Risk Factors for Death from Visceral Leishmaniasis in an Urban Area of Brazil. PLoS Negl Trop Dis. 2015;9: e0003982. doi:10.1371/journal.pntd.0003982

17. Cota GF, de Sousa MR, de Mendonça ALP, Patrocinio A, Assunção LS, de Faria SR, et al. Leishmania-HIV Co-infection: Clinical Presentation and Outcomes in an Urban Area in Brazil. Debrabant A, editor. PLoS Negl Trop Dis. 2014;8: e2816. doi:10.1371/journal.pntd.0002816

18. Souza GF de, Biscione F, Greco DB, Rabello A. Slow clinical improvement after treatment initiation in Leishmania/HIV coinfected patients. Rev Soc Bras Med Trop. 2012;45: 147–150. doi:10.1590/S0037-86822012000200001

19. Sousa JM dos S, Ramalho WM, De Melo MA. Demographic and clinical characterization of human visceral leishmaniasis in the State of Pernambuco, Brazil between 2006 and 2015. Rev Soc Bras Med Trop. 2018;51: 622–630. doi:10.1590/0037-8682-0047-2018

20. Soares VYR, Lúcio Filho CEP, Carvalho LIM de, Silva AMM de M e, Eulálio KD. Clinical and epidemiological analysis of patients with HIV/AIDS admitted to a reference hospital in the northeast region of Brazil. Rev Inst Med Trop Sao Paulo. 2008;50: 327–332. doi:10.1590/S0036-46652008000600003

21. Lima ÁLM, de Lima ID, Coutinho JFV, de Sousa ÚPST, Rodrigues MAG, Wilson ME, et al. Changing epidemiology of visceral leishmaniasis in northeastern Brazil: A 25-year follow-up of an urban outbreak. Trans R Soc Trop Med Hyg. 2017;111: 440–447. doi:10.1093/trstmh/trx080

22. Lima ID, Lima ALM, Mendes-Aguiar C de O, Coutinho JFV, Wilson ME, Pearson RD, et al. Changing demographics of visceral leishmaniasis in northeast Brazil: Lessons for the future. PLoS Negl Trop Dis. 2018;12: 1–16. doi:10.1371/journal.pntd.0006164

23. Santos GDO, De Jesus NPS, Cerqueira-Braz JV, Santos VS, De Lemos LMD. Prevalence of HIV and associated factors among visceral leishmaniasis cases in an endemic area of Northeast Brazil. Rev Soc Bras Med Trop. 2019;52: 1–4. doi:10.1590/0037-8682-0257-2018

24. Albuquerque LCP de, Mendonça IR, Cardoso PN, Baldaçara LR, Borges MRMM, Borges J da C, et al. HIV/AIDS-related visceral leishmaniasis: a clinical and epidemiological description of visceral leishmaniasis in northern Brazil. Rev Soc Bras Med Trop. 2014;47: 38–46. doi:10.1590/0037-8682-0180-2013

25. Secretaria de Saúde do Estado da Bahia. Informe Epidemiológico de Leishmaniose Visceral, Bahia. 2018.

26. Secretaria Municipal de Saúde de Salvador. Boletim Epidemiológico de AIDS: Situação Epidemiológica da AIDS em Salvador, Bahia. 2019.

27. Charlab R, Barral-Netto M, Valenzuela JG, Barral A, Caldas A, Honda E, et al. Human immune response to sand fly salivary gland antigens: a useful epidemiological marker? Am J Trop Med Hyg. 2000;62: 740–745. doi:10.4269/ajtmh.2000.62.740

28. Souza AP, Soto M, Costa JML, Boaventura VS, de Oliveira CI, Cristal JR, et al. Towards a More Precise Serological Diagnosis of Human Tegumentary Leishmaniasis Using Leishmania Recombinant Proteins. Zamboni DS, editor. PLoS One. 2013;8: e66110. doi:10.1371/journal.pone.0066110

29. Soto M, Corvo L, Garde E, Ramírez L, Iniesta V, Bonay P, et al. Coadministration of the Three Antigenic Leishmania infantum Poly (A) Binding Proteins as a DNA Vaccine Induces Protection against Leishmania major Infection in BALB/c Mice. Melby PC, editor. PLoS Negl Trop Dis. 2015;9: e0003751. doi:10.1371/journal.pntd.0003751

30. Barreto CC, Nishyia A, Araújo L V., Ferreira JE, Busch MP, Sabino EC. Trends in antiretroviral drug resistance and clade distributions among HIV-1-infected blood donors in Sao Paulo, Brazil. J Acquir Immune Defic Syndr. 2006;41: 338– 341. doi:10.1097/01.qai.0000199097.88344.50

31. Kozal MJ, Shah N, Shen N, Yang R, Fucini R, Merigan TC, et al. Extensive polymorphisms observed in HIV–1 clade B protease gene using high–density oligonucleotide arrays. Nat Med. 1996;2: 753–759. doi:10.1038/nm0796-753

32. Pieniazek D, Peralta JM, Ferreira JA, Krebs JW, Owen SM, Sion FS, et al. Identification of mixed HIV-1/HIV-2 infections in Brazil by polymerase chain reaction. AIDS. 1991;5: 1293–1300. doi:10.1097/00002030-199111000-00002

33. Frenkel LM, Wagner LE, Atwood SM, Cummins TJ, Dewhurst S. Specific, sensitive, and rapid assay for human immunodeficiency virus type 1 pol mutations associated with resistance to zidovudine and didanosine. J Clin Microbiol. 1995;33: 342–347. doi:10.1128/jcm.33.2.342-347.1995

34. Janini LM, Pieniazek D, Peralta JM, Schechter M, Tanuri A, Vicente ACP, et al. Identification of single and dual infections with distinct subtypes of human immunodeficiency virus type 1 by using restriction fragment length polymorphism analysis. Virus Genes. 1996;13: 69–81. doi:10.1007/BF00576981

35. Schultz A-K, Zhang M, Bulla I, Leitner T, Korber B, Morgenstern B, et al. jpHMM: Improving the reliability of recombination prediction in HIV-1. Nucleic Acids Res. 2009;37: W647–W651. doi:10.1093/nar/gkp371

36. Guindon S, Dufayard J-F, Lefort V, Anisimova M, Hordijk W, Gascuel O. New Algorithms and Methods to Estimate Maximum-Likelihood Phylogenies: Assessing the Performance of PhyML 3.0. Syst Biol. 2010;59: 307–321. doi:10.1093/sysbio/syq010

37. Vivariniáislan de C, Calegari-Silva TC, Saliba AM, Boaventura VS, França-Costa J, Khouri R, et al. Systems Approach Reveals Nuclear Factor Erythroid 2-Related Factor 2/Protein Kinase R Crosstalk in Human Cutaneous Leishmaniasis. Front Immunol. 2017;8: 1–18. doi:10.3389/fimmu.2017.01127

38. Subramanian K, Dierckx T, Khouri R, Menezes SM, Kagdi H, Taylor GP, et al. Decreased RORC expression and downstream signaling in HTLV-1-associated adult T-cell lymphoma/leukemia uncovers an antiproliferative IL17 link: A potential target for immunotherapy? Int J Cancer. 2019;144: 1664–1675. doi:10.1002/ijc.31922

39. Delgobo M, Mendes DAGB, Kozlova E, Rocha EL, Rodrigues-Luiz GF, Mascarin L, et al. An evolutionary recent IFN/IL-6/CEBP axis is linked to monocyte expansion and tuberculosis severity in humans. Elife. 2019;8: 1–32. doi:10.7554/eLife.47013

40. Rotger M, Dang KK, Fellay J, Heinzen EL, Feng S, Descombes P, et al. Genome-Wide mRNA Expression Correlates of Viral Control in CD4+ T-Cells from HIV-1-Infected Individuals. Emerman M, editor. PLoS Pathog. 2010;6: e1000781. doi:10.1371/journal.ppat.1000781

41. Gardinassi LG, Garcia GR, Costa CHN, Costa Silva V, de Miranda Santos IKF. Blood Transcriptional Profiling Reveals Immunological Signatures of Distinct States of Infection of Humans with Leishmania infantum. Acosta-Serrano A, editor. PLoS Negl Trop Dis. 2016;10: e0005123. doi:10.1371/journal.pntd.0005123

42. Zacarias DA, Rolão N, de Pinho FA, Sene I, Silva JC, Pereira TC, et al. Causes and consequences of higher Leishmania infantum burden in patients with kala-azar: a study of 625 patients. Trop Med Int Heal. 2017;22: 679–687. doi:10.1111/tmi.12877

43. Bogdan C, Donhauser N, Döring R, Röllinghoff M, Diefenbach A, Rittig MG. Fibroblasts as host cells in latent leishmaniosis. J Exp Med. 2000;191: 2121–2129. doi:10.1084/jem.191.12.2121

44. Paduch K, Debus A, Rai B, Schleicher U, Bogdan C. Resolution of Cutaneous Leishmaniasis and Persistence of Leishmania major in the Absence of Arginase 1. J Immunol. 2019;202: 1453–1464. doi:10.4049/jimmunol.1801249

45. Seyed N, Peters NC, Rafati S. Translating observations from Leishmanization into non-living vaccines: The potential of dendritic cell-based vaccination strategies against Leishmania. Front Immunol. 2018;9: 1–10. doi:10.3389/fimmu.2018.01227

46. Caldas A, Favali C, Aquino D, Vinhas V, van Weyenbergh J, Brodskyn C, et al. Balance of IL-10 and interferon-γ plasma levels in human visceral leishmaniasis: Implications in the pathogenesis. BMC Infect Dis. 2005;5: 1–9. doi:10.1186/1471-2334-5-113

47. Frade AF, Oliveira LC de, Costa DL, Costa CHN, Aquino D, Van Weyenbergh J, et al. TGFB1 and IL8 gene polymorphisms and susceptibility to visceral leishmaniasis. Infect Genet Evol. 2011;11: 912–916. doi:10.1016/j.meegid.2011.02.014

48. Carneiro MW, Fukutani KF, Andrade BB, Curvelo RP, Cristal JR, Carvalho AM, et al. Gene Expression Profile of High IFN-γ Producers Stimulated with Leishmania braziliensis Identifies Genes Associated with Cutaneous Leishmaniasis. McDowell MA, editor. PLoS Negl Trop Dis. 2016;10: e0005116. doi:10.1371/journal.pntd.0005116

49. Peruhype-Magalhães V, Martins-Filho OA, Prata A, Silva LDA, Rabello A, Teixeira-Carvalho A, et al. Mixed inflammatory/regulatory cytokine profile marked by simultaneous raise of interferon-γ and interleukin-10 and low frequency of tumour necrosis factor-α+ monocytes are hallmarks of active human visceral Leishmaniasis due to Leishmania chagasi infectio. Clin Exp Immunol. 2006;146: 124–132. doi:10.1111/j.1365-2249.2006.03171.x

50. Botana L, Ibarra-Meneses AV, Sánchez C, Castro A, Martin JVS, Molina L, et al. Asymptomatic immune responders to Leishmania among HIV positive patients. PLoS Negl Trop Dis. 2019;13: 1–14. doi:10.1371/journal.pntd.0007461

51. Horrillo L, Castro A, Matía B, Molina L, García-Martínez J, Jaqueti J, et al. Clinical aspects of visceral leishmaniasis caused by L. infantum in adults. Ten years of experience of the largest outbreak in Europe: what have we learned? Parasit Vectors. 2019;12: 359. doi:10.1186/s13071-019-3628-z

52. Ana Nilce S M, Adolfo G, Romero S, Samantha YOB, Lindoso AL, Elisa C, et al. Premature deaths by visceral leishmaniasis in Brazil investigated through a cohort study : A challenging opportunity ? PLoS Negl Trop Dis. 2019;13: 1–18. doi:https://doi.org/10.1371/journal.pntd.0007841

53. Van Griensven J, van Henten S, Mengesha B, Kassa M, Adem E, Seid ME, et al. Longitudinal evaluation of asymptomatic Leishmania infection in HIV-infected individuals in North-West Ethiopia: A pilot study. PLoS Negl Trop Dis. 2019;13: 30–45. doi:10.1371/journal.pntd.0007765

54. Diro E, Edwards T, Ritmeijer K, Fikre H, Abongomera C, Kibret A, et al. Long term outcomes and prognostics of visceral leishmaniasis in hiv infected patients with use of pentamidine as secondary prophylaxis based on CD4 level: A prospective cohort study in Ethiopia. PLoS Negl Trop Dis. 2019;13: 1–17. doi:10.1371/journal.pntd.0007132

55. Prestes-Carneiro LE, Spir PRN, Fontanesi M, Pereira Garcia KG, Silva FA Da, Flores EF, et al. Unusual manifestations of visceral leishmaniasis in children: A case series and its spatial dispersion in the western region of São Paulo state, Brazil. BMC Infect Dis. 2019;19: 1–9. doi:10.1186/s12879-018-3652-1

56. Fontoura IG, Barbosa DS, de Andrade Paes AM, Santos FS, Neto MS, Fontoura VM, et al. Epidemiological, clinical and laboratory aspects of human visceral leishmaniasis (HVL) associated with human immunodeficiency virus (HIV) coinfection: a systematic review – CORRIGENDUM. Parasitology. 2018;145: 1819–1819. doi:10.1017/S0031182018001166

57. Rezaei Z, Sarkari B, Dehghani M, Layegh Gigloo A, Afrashteh M. High frequency of subclinical Leishmania infection among HIV-infected patients living in the endemic areas of visceral leishmaniasis in Fars province, southern Iran. Parasitol Res. 2018;117: 2591–2595. doi:10.1007/s00436-018-5949-9

58. Viana GM de C, da Silva MACN, Garcia JV de S, Guimarães HD, Arcos Júnior GF, Santos AVA, et al. Epidemiological profile of patients co-infected with visceral leishmaniasis and HIV/AIDS in northeast, Brazil. Rev Soc Bras Med Trop. 2017;50: 613–620. doi:10.1590/0037-8682-0494-2017

59. Orsini M, Canela JR, Disch J, Maciel F, Greco D, Toledo A, et al. High frequency of asymptomatic Leishmania spp. infection among HIV-infected patients living in endemic areas for visceral leishmaniasis in Brazil. Trans R Soc Trop Med Hyg. 2012;106: 283–288. doi:10.1016/j.trstmh.2012.01.008

60. Fukutani KF, Figueiredo V, Celes FS, Cristal JR, Barral A, Barral-Netto M, et al. Serological survey of Leishmaniainfection in blood donors in Salvador, Northeastern Brazil. BMC Infect Dis. 2014;14: 422. doi:10.1186/1471-2334-14-422

61. Oryan A, Akbari M. Worldwide risk factors in leishmaniasis. Asian Pac J Trop Med. 2016;9: 925–932. doi:10.1016/j.apjtm.2016.06.021

62. Grande E, Zucchetto A, Suligoi B, Grippo F, Pappagallo M, Virdone S, et al. Multiple cause-of-death data among people with AIDS in Italy: A nationwide cross-sectional study. Popul Health Metr. 2017;15: 1–12. doi:10.1186/s12963-017-0135-3

63. Bruhn FRP, Morais MHF, Bruhn NCP, Cardoso DL, Ferreira F, Rocha CMBM. Human visceral leishmaniasis: Factors associated with deaths in Belo Horizonte, Minas Gerais state, Brazil from 2006 to 2013. Epidemiol Infect. 2018;146: 565–570. doi:10.1017/S0950268818000109

64. Guedes DL, Medeiros Z, Da Silva ED, De Vasconcelos AVM, Da Silva MS, Da Silva MAL, et al. Visceral leishmaniasis in hospitalized HIV-infected patients in Pernambuco, Brazil. Am J Trop Med Hyg. 2018;99: 1541–1546. doi:10.4269/ajtmh.17-0787

65. Braga L de S, Navasconi TR, Leatte EP, Skraba CM, Silveira TGV, Ribas-Silva RC. Presence of anti-leishmania (viannia) braziliensis antibodies in blood donors in the west-central region of the state of Paraná, Brazil. Rev Soc Bras Med Trop. 2015;48: 622–625. doi:10.1590/0037-8682-0043-2015

66. Monteiro DCS, Sousa AQ, Lima DM, Fontes RM, Praciano CC, Frutuoso MS, et al. Leishmania infantum Infection in Blood Donors, Northeastern Brazil. Emerg Infect Dis. 2016;22: 739–740. doi:10.3201/eid2204.150065

67. le Fichoux Y, Quaranta JF, Aufeuvre JP, Lelievre A, Marty P, Suffia I, et al. Occurrence of Leishmania infantum parasitemia in asymptomatic blood donors living in an area of endemicity in southern France. J Clin Microbiol. 1999;37: 1953–7.

68. de Oliveira França A, Pompilio MA, Pontes ERJC, de Oliveira MP, Pereira LOR, Lima RB, et al. Leishmania infection in blood donors: A new challenge in leishmaniasis transmission? PLoS One. 2018;13: 1–13. doi:10.1371/journal.pone.0198199

69. Khouri R, Vandamme AM. Virus genetic variability involvement in transmissibility of HIV-1 immune activation and disease progression. Future Virol. 2015;10: 1259–1262. doi:10.2217/fvl.15.96

70. Araujo AF, Brites C, Monteiro-Cunha J, Santos LA, Galvao–Castro B, Alcantara LCJ. Lower Prevalence of Human Immunodeficiency Virus Type 1 Brazilian Subtype B Found in Northeastern Brazil with Slower Progression to AIDS. AIDS Res Hum Retroviruses. 2010;26: 1249–1254. doi:10.1089/aid.2010.0068

71. Monteiro-Cunha JP, Araujo AF, Santos E, Galvao-Castro B, Alcantara LCJ. Lack of High-Level Resistance Mutations in HIV Type 1 BF Recombinant Strains Circulating in Northeast Brazil. AIDS Res Hum Retroviruses. 2011;27: 623–631. doi:10.1089/aid.2010.0126

72. Santos LA, Monteiro-Cunha JP, Araújo AF, Brites C, Galvão-Castro B, Alcantara LCJ. Detection of Distinct Human Immunodeficiency Virus Type 1 Circulating Recombinant Forms in Northeast Brazil. J Med Virol. 2011;83: 2066–2072. doi:10.1002/jmv

73. Chavez HH, Tran T-A, Dembele B, Nasreddine N, Lambotte O, Gubler B, et al. Lack of Evidence for Prolonged Double-Long Terminal Repeat Episomal HIV DNA Stability In Vivo. JAIDS J Acquir Immune Defic Syndr. 2007;45: 247–249. doi:10.1097/QAI.0b013e3180415dc2

74. Santos EDS, Araújo AF, Galvão-Castro B, Alcantara LCJ. Diversidade genética do vírus da imunodeficiência humana tipo 1 (HIV-1) em mulheres infectadas de uma cidade do nordeste do Brasil. Rev Bras Ginecol e Obs. 2009;31: 609–614. doi:10.1590/S0100-72032009001200006

75. Monteiro JP, Alcantara LCJ, de Oliveira T, Oliveira AM, Melo MAG, Brites C, et al. Genetic variability of human immunodeficiency virus-1 in Bahia state, Northeast, Brazil: High diversity of HIV genotypes. J Med Virol. 2009;81: 391– 399. doi:10.1002/jmv.21414

76. Hoffmann M, Pantazis N, Martin GE, Hickling S, Hurst J, Meyerowitz J, et al. Exhaustion of Activated CD8 T Cells Predicts Disease Progression in Primary HIV-1 Infection. Wanstrom R, editor. PLOS Pathog. 2016;12: e1005661. doi:10.1371/journal.ppat.1005661

77. Ipp H, Zemlin AE, Erasmus RT, Glashoff RH. Role of inflammation in HIV-1 disease progression and prognosis. Crit Rev Clin Lab Sci. 2014;51: 98–111. doi:10.3109/10408363.2013.865702

78. Utay NS, Hunt PW. Role of immune activation in progression to AIDS. Curr Opin HIV AIDS. 2016;11: 131–137. doi:10.1097/COH.0000000000000242

79. Kedzierska K, Crowe SM. Cytokines and HIV-1: Interactions and Clinical Implications. Antivir Chem Chemother. 2001;12: 133–150. doi:10.1177/095632020101200301

80. Kenway-Lynch CS, Das A, Lackner AA, Pahar B. Cytokine/Chemokine Responses in Activated CD4+ and CD8+ T Cells Isolated from Peripheral Blood, Bone Marrow, and Axillary Lymph Nodes during Acute Simian Immunodeficiency Virus Infection. J Virol. 2014;88: 9442–9457. doi:10.1128/JVI.00774-14

81. Mehla R. Chemokine Deregulation in HIV Infection: Role of Interferon Gamma Induced Th1-Chemokine Signaling. J Clin Cell Immunol. 2013;04. doi:10.4172/2155-9899.S7-004

82. Utay NS, Douek DC. Interferons and HIV Infection: The Good, the Bad, and the Ugly. Pathog Immun. 2016;1: 107. doi:10.20411/pai.v1i1.125

83. Accornero P, Radrizzani M, Delia D, Gerosa F, Kurrle R, Colombo MP. Differential Susceptibility to HIV-GP120–Sensitized Apoptosis in CD4+ T-Cell Clones With Different T-Helper Phenotypes: Role of CD95/CD95L Interactions. Blood. 1997;89: 558–569. doi:10.1182/blood.V89.2.558

84. Cummins NW, Badley AD. Mechanisms of HIV-associated lymphocyte apoptosis: 2010. Cell Death Dis. 2010;1: e99–e99. doi:10.1038/cddis.2010.77

85. Costa ASA, Costa GC, Aquino DMC de, Mendonça VRR de, Barral A, Barral-Netto M, et al. Cytokines and visceral leishmaniasis: a comparison of plasma cytokine profiles between the clinical forms of visceral leishmaniasis. Mem Inst Oswaldo Cruz. 2012;107: 735–739. doi:10.1590/S0074-02762012000600005

86. Rodrigues V, Cordeiro-da-Silva A, Laforge M, Silvestre R, Estaquier J. Regulation of immunity during visceral Leishmania infection. Parasit Vectors. 2016;9: 118. doi:10.1186/s13071-016-1412-x

87. Sun H, Kim D, Li X, Kiselinova M, Ouyang Z, Vandekerckhove L, et al. Th1/17 Polarization of CD4 T Cells Supports HIV-1 Persistence during Antiretroviral Therapy. Silvestri G, editor. J Virol. 2015;89: 11284–11293. doi:10.1128/JVI.01595-15

88. Khouri R, Silva-Santos G, Dierckx T, Menezes SM, Decanine D, Theys K, et al. A genetic IFN/STAT1/FAS axis determines CD4 T stem cell memory levels and apoptosis in healthy controls and Adult T-cell Leukemia patients. Oncoimmunology. 2018;7: e1426423. doi:10.1080/2162402X.2018.1426423

89. Farre L, Bittencourt AL, Silva-Santos G, Almeida A, Silva AC, Decanine D, et al. Fas-670 promoter polymorphism is associated to susceptibility, clinical presentation, and survival in adult T cell leukemia. J Leukoc Biol. 2008;83: 220– 222. doi:10.1189/jlb.0407198

